# Analysis of Potential Risk Factors of COVID-19 Based on Variants: Omicron, Delta, and Alpha

**DOI:** 10.1101/2023.09.26.23295911

**Authors:** Dharshini Kannan, Sreenidhi Muppiri, Maren Reid

## Abstract

The coronavirus disease 2019 (COVID-19) pandemic has changed which affects the risk of COVID-19 infection for specific subgroups. We focused on the subgroups based on the factors (sex, age, and vaccination) and COVID-19 strains (Alpha, Delta, and Omicron). Past studies focused on analyzing these factors based on one geographic region or one COVID-19 strain. Therefore, there is a need to understand these factors’ association with risk of COVID-19 infection through analyzing data from various geographic regions and strains. The association between COVID-19 strains and the factors was assessed through chi-square test and odds ratio tests. Sex, vaccination, age had a significant association with testing positive for the COVID-19 strains of interest in most geographies. The biggest difference was unvaccinated individuals have 3.14 higher odds of getting Alpha than vaccinated individuals in Canada. These findings provide insights into the groups that are more susceptible to contracting specific strains of COVID-19.

**Summary:** This manuscript offers a broad examination of key factors that may impact the risk of COVID-19 infection across various geographic regions. Results provide evidence about potential risk factors for COVID-19 infection around the world for certain sub-groups as COVID-19 mutates.

## Introduction

The coronavirus disease 2019 (COVID-19) began to spread around the world at the end of 2019, causing severe upper-respiratory disease and death, overwhelming hospitals, draining emergency resources, and inciting fear and panic. On March 11, 2020, the World Health Organization (WHO) declared COVID-19 as a pandemic.^1^ As of May 5, 2023, WHO has declared that COVID-19 is no longer a public health emergency of international concern (PHEIC).^1^ Between 2020 and 2023, the virus mutated multiple times and various variants emerged. The variants of concern (VOCs) are Delta (B.1.617.2), Beta (B.1.351), Omicron (B.1.1.529), and Alpha (B.1.1.7) as of 2023.^2^ Even with mitigating measures such as vaccination, the spread of COVID-19 has not significantly decreased when it comes to new COVID-19 variants.^3^ Thus, there is a need to understand further the risk factors for COVID-19 and its past variants to better control the spread of COVID-19.

Epidemiological studies identifying the risk factors for infection by certain variants are essential to understanding the spread of COVID-19 and improving patient care management. A number of studies examined risk factors such as age, sex, and vaccination. In Europe, there is no material association between COVID-19 infection by age or sex.^4^ Studies from the US and Spain seem to suggest a competing result: 52% and a higher percentage of patients hospitalized with COVID-19 were males.^5^ Vaccination rates and uptake are also differential by population; data from South Africa suggest that in 2021, 29.2% of the adult population reported being hesitant about receiving a COVID-19 vaccine.^6^ Based on the study, South African population is less likely to be vaccinated, increasing the incidence of COVID-19 for a proportion of their population. In contrast, Israel reported higher levels of vaccine uptake, but data also showed a seemingly increased incidence of COVID-19 for individuals who had a significant amount of time elapsed after the second vaccination dose of the Pfizer-BioNTech BNT162b2 mRNA vaccine.^7^ Even so, there is abundant evidence demonstrating that overall, vaccination protects against hospitalization due to COVID-19 complications.^8^ Therefore, the discordant findings across studies from different geographies and populations suggest that more investigation is necessary to fully understand the nuanced and/or differential impact of sex, age, and vaccination on the risk of COVID-19.

A comprehensive review of the risk factors of COVID-19 infection in countries across the world is essential to understanding gaps in COVID-19 infection patterns, enhancing preventive care for COVID-19 patients, and improving care management for COVID-19 patients. To this end, this study assessed associations between COVID-19 strain and risk factors, including age, sex, and vaccination status, through a modified, meta-analysis approach.

## Methods

We used a meta-analysis approach to conduct a summary analysis of key studies representing diverse geographies and populations to identify key demographic factors associated with strain-specific COVID-19 infection. Though this analysis is not strictly a meta-analysis, we chose to align the methodology as closely as possible to the Preferred Reporting Items for Systematic Reviews and Meta-Analyses (PRISMA) guidelines and checklist.

### Search Strategy

We searched two databases - PubMed and the Incidence and Prevalence Database (IPD)- for studies published between December 1, 2020 and January 5, 2023 to identify relevant COVID-19 publications which reported demographics, symptoms, and vaccination status of people testing positive for COVID-19. The keywords and Medical Subject Headings (MeSH) terms selected included “COVID-19,” “SARS-CoV-2,” and “Novel coronavirus 2019.” We performed the last search on January 5, 2023.

### Eligibility criteria and study selection

#### Inclusion criteria for screening

Our study methodology started with screening for articles using only study titles and abstracts. Those deemed eligible for analysis consideration were further scrutinized in a second step for inclusion into the analysis set.

We screened peer-reviewed and non-peer-reviewed studies for papers analyzing populations with confirmed SARS-CoV-2 via positive polymerase chain reaction (PCR) test, reverse transcriptase polymerase chain reaction (RT-PCR), molecular testing (laboratory-based or qualitative antigen test), or respiratory samples (nasopharyngeal swabs, sputum, and bronchoalveolar lavage fluid). Further, studies confirmed infection strain via whole-genome sequencing (WGS) or S gene target failure (SGTF). Selected articles also included variables on demographics, outcomes, risk factors, vaccination, clinical manifestations, and symptoms. Finally, eligible studies were not limited by language of publication; we used Google Translate tools to translate non-English articles to English. We excluded the studies if they focused on a certain age group or other subpopulation; geographic regions had a population of less than one million; subjects weren’t hospitalized for COVID-19 or only considered patients who had the same outcome. The publication types excluded were reprints and studies such as letters, correspondence, and comments. Articles with irrelevant clinical study focus (such as animal data and care management) and nonclinical study focus (such as diagnostic techniques) were also excluded.

#### Inclusion criteria

Studies that passed the initial screening were further filtered to identify papers with the aligned variables and sample size required for the primary analysis. For the primary analysis, studies needed to report the vaccination status, sex, or gender of each subject in the study. The investigators extracted ecological data. The final study sample included studies that all contained data on confirmed SARS-CoV-2 diagnosis and strain in addition to data on vaccination, demographics, including sex and age, and symptoms of fever and pneumonia.

### Data extraction

Two investigators independently worked on title/abstract, full-text screening, and data extraction on a shared data extraction on a shared extraction form. Investigators reached initial agreement across all information extracted for most of the studies, but they discussed any disagreements until they reached a consensus. The screening process produced a total of 10 studies containing data on 128,841 subjects (assuming these sources didn’t repeat any individual’s data within and between these sources). The average number of people per source was 12,884 for this research. The minimum total sample size of a source included in this study is 65; the maximum number of people in the source included in this study is 78,166.

Of the 144 studies identified, 10 studies were selected for analysis (Figure 1). We extracted data for vaccination status, symptoms of pneumonia and fever, and demographic characteristics. We categorized sex as male and female; age as ≥ 60 and < 60 years old; vaccination categories as no vaccination (zero vaccine doses) or any vaccination (one or more vaccine doses). Note that 9 studies did not report sex or account for non-binary, gender-questioning, or transgender status of their participants (1 didn’t report sex and 8 didn’t account of other genders than male and female). 7 studies did not report age or include subjects aligning with the age range of interest for our analysis, and 6 studies did not report vaccination status for their analysis population. In these cases, we considered these fields missing.

**Figure 1.**
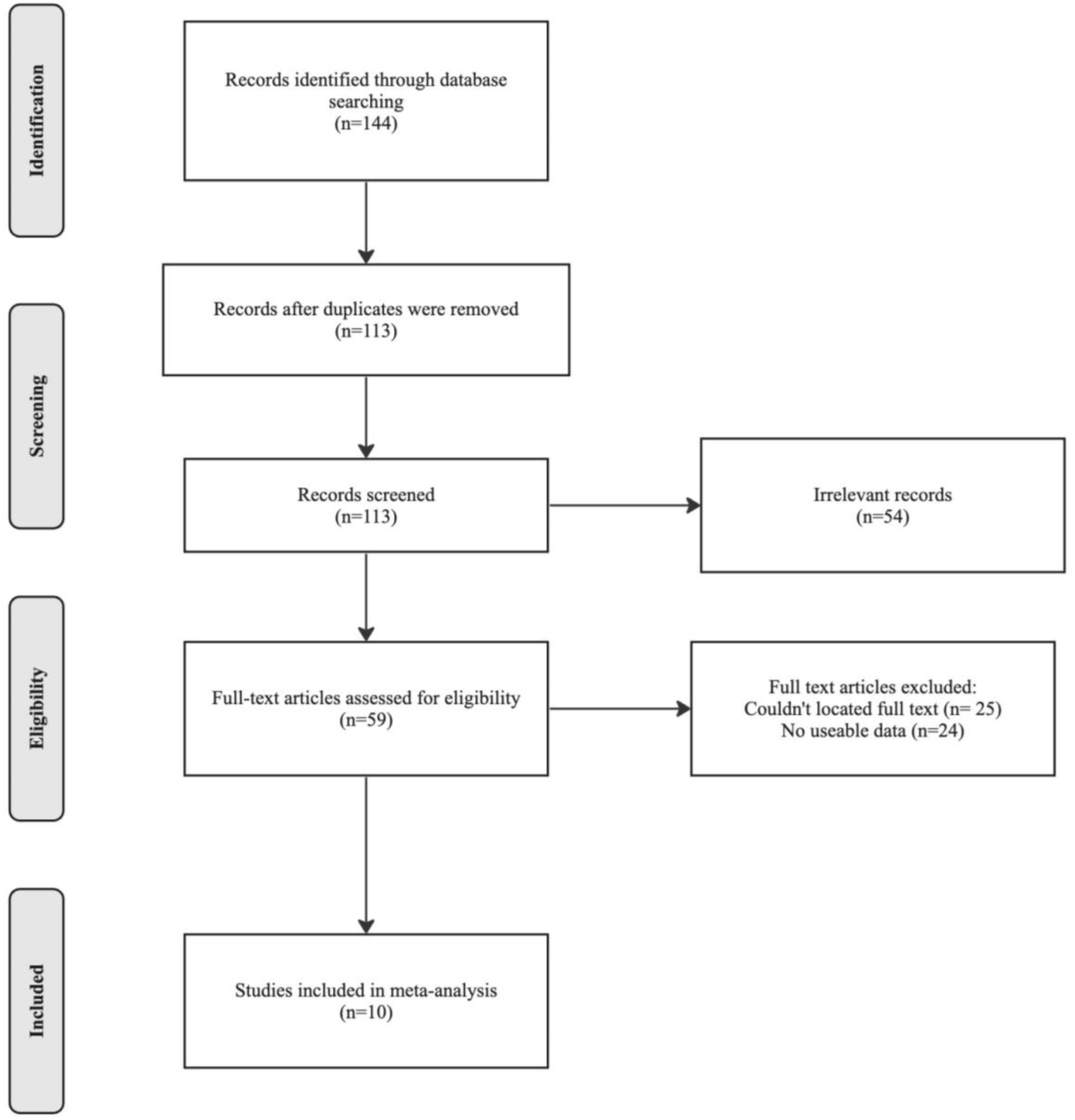
Flow diagram of the article screening process.

Since fever is a generalized symptom for a variety of illnesses, when possible, studies omitted fever due to pneumonia by using the scoring system proposed by the Japanese Respiratory System (JRS). Non-pneumonia fevers were therefore included in the studies if subjects presented with a fever which was greater than or equal to 37.0 degrees Celsius. This study did not consider symptom combinations pertaining to pneumonia and fever due to a lack of studies recording data on the number of COVID-19 patients with both.

The COVID-19 strains of interest for this study were those categorized as variants of concern as of 2023 - Omicron, Delta, and Alpha (see Table 1 for virology phenotypes). Beta is excluded from our analysis due to lack of studies meeting the eligibility criteria. We categorized studies by the strain reported in the analysis population. If studies contained more than one COVID-19 strain, we binned those studies as a "multiple strains" category. The final, analysis set of studies summarized in this analysis are from South Africa, Canada, China, Japan, Bahrain, Spain, England, and Singapore.

**Table 1.** Virology Phenotype of COVID-19 Strains (Omicron, Delta, Alpha)

|  | COVID-19 Strains Included in This Study |  |  |
| --- | --- | --- | --- |
|  | Omicron (B.1.1.529) | Delta (B.1.617.2) | Alpha (B.1.1.7) |
| Sublineage of the<br>COVID-19 Variant | BA.1 | AY.25 | B.1.1.7 |
|  | BA.2 | AY.27 |  |
|  | BA.3 | Delta1 |  |
|  | BA.4 | Delta2 |  |
|  | BA.5 | AY.4.2 |  |
|  | B.1.1.529 | AY.4 |  |
|  |  | AY.6 |  |
|  |  | B.1.617.2 |  |

### Statistical Analysis

Using *X*^2^ test of association, we conducted a bivariate assessment for each factor-COVID-19 strain pair by study for each study in the sample. P-values of <0.05 were considered statistically significant. To estimate the strength and direction of the association between covariates of interest and COVID-19 strains, we also report odds ratios (OR) and 95% confidence intervals. To calculate the OR, we subsetted the data on the characteristics of interest from the analysis population. This study used the odds ratio (calculated by median-unbiased estimate) with a 95% confidence interval (calculated by mid-p exact confidence interval) to identify statistically significant odds between unvaccinated and vaccinated, male and female, and sixty plus and less than sixty groups using the Epitools package in R.

## Results

A total of 128,841 patients diagnosed with COVID-19 were identified and included in the analysis sample (Table 2). Of these patients, 46,240 (35.9%), 46,335 (36.0%), and 36,266 (28.1%) were diagnosed with Omicron, Delta, and Alpha COVID-19 strain, respectively. A higher proportion of the sample infected with Delta were vaccinated compared to those diagnosed with Alpha [18.9% vs 7.3%]. Similarly, people testing positive for Omicron had a higher proportion of vaccination than Delta patients [26.4% vs. 18.9%] and tended to be older than people diagnosed with Delta or Alpha.

**Table 2.** Meta – Sample: Sample size of selected studies and summary of demographics and vaccination data.

|  |  | South Africa |  | China |  | Japan |  |
| --- | --- | --- | --- | --- | --- | --- | --- |
|  |  | (Abdullah et al., 2022)<sup>28</sup> |  | (Bi et al., 2022)<sup>29</sup> |  | (Ito et al., 2022)<sup>30</sup> |  |
|  |  | N | Value (%) | N | Value (%) | N | Value (%) |
| Total Number of Patients |  | 4,428 | 100.0 | 65 | 100.0 | 231 | 100.0 |
| COVID-19 Strain(s) |  |  |  |  |  |  |  |
| Omicron |  | 4,428 | 100.0 | 65 | 100.0 | 231 | 100.0 |
| Delta |  | * | * | * | * | * | * |
| Alpha |  | * | * | * | * | * | * |
| Sex |  |  |  |  |  |  |  |
| Female |  | * | * | 30 | 46.2 | 117 | 50.6 |
| *Omicron* |  | * | * | 30 | 46.2 | 117 | 50.6 |
| *Delta* |  | * | * | * | * | * | * |
| *Alpha* |  | * | * | * | * | * | * |
| Male |  | * | * | 35 | 53.8 | 116 | 50.2 |
| *Omicron* |  | * | * | 35 | 53.8 | 116 | 50.2 |
| *Delta* |  | * | * | * | * | * | * |
| *Alpha* |  | * | * | * | * | * | * |

Note. \*=Data not reported

Table 2 (Continued)
|  | **Canada** |  | **Bahrain** |  | **Japan** |  |
| --- | --- | --- | --- | --- | --- | --- |
|  | **(Ulloa et al., 2022)<sup>31</sup>** |  | **(Kumar et al., 2022)<sup>32</sup>** |  | **(Miyashita et al., 2022)<sup>33</sup>** |  |
|  | N | Value (%) | N | Value (%) | N | Value (%) |
| Total Number of Patients | 78,166 | 100.0 | 1,428 | 100.0 | 335 | 100.0 |
| COVID-19 Strain(s) |  |  |  |  |  |  |
| Omicron | 41,216 | 52.7 | * | * | * | * |
| Delta | 36,950 | 47.3 | 636 | 44.5 | * | * |
| Alpha | * | * | 792 | 55.5 | 335 | 100.0 |
| Sex |  |  |  |  |  |  |
| Female | 38,822 | 49.7 | 462 | 32.4 | 108 | 32.2 |
| *Omicron* | 20,562 | 26.3 | * | * | * | * |
| *Delta* | 18,260 | 23.4 | 137 | 9.6 | * | * |
| *Alpha* | * | * | 325 | 22.8 | 108 | 32.2 |
| Male | 39,234 | 50.2 | 866 | 60.6 | 227 | 67.8 |
| *Omicron* | 20,614 | 26.4 | * | * | * | * |
| *Delta* | 18,620 | 23.8 | 399 | 27.9 | * | * |
| *Alpha* | * | * | 467 | 32.7 | 227 | 67.8 |

Note. \*=Data not reported

Table 2 (Continued)
|  | **Spain** |  | **England** |  | **Singapore** |  |
| --- | --- | --- | --- | --- | --- | --- |
|  | **(Martínez-García et al.,**<br>**2021)<sup>34</sup>** |  | **(Twohig et al.,**<br>**2022)<sup>35</sup>** |  | **(Ong et al., 2022)<sup>36</sup>** |  |
|  | N | Value (%) | N | Value (%) | N | Value (%) |
| Total Number of Patients | 426 | 100.0 | 43,338 | 100.00% | 124 | 100.0 |
| COVID-19 Strain(s) |  |  |  |  |  |  |
| Omicron | * | * | * | * | * | * |
| Delta | * | * | 8,682 | 20.0 | 67 | 54.0 |
| Alpha | 426 | 100.0 | 34,656 | 80.0 | 57 | 46.0 |
| Sex |  |  |  |  |  |  |
| Female | 251 | 58.9 | 22,162 | 51.1 | 55 | 44.4 |
| *Omicron* | * | * | * | * | * | * |
| *Delta* | * | * | 4,249 | 9.8 | 33 | 26.6 |
| *Alpha* | 251 | 58.9 | 17,913 | 41.3 | 22 | 17.7 |
| Male | 175 | 41.1 | 21,176 | 48.9 | 69 | 55.6 |
| *Omicron* | * | * | * | * | * | * |
| *Delta* | * | * | 4,433 | 10.2 | 34 | 27.4 |
| *Alpha* | 175 | 41.1 | 16,743 | 38.6 | 35 | 28.2 |

Note. \*=Data not reported

Table 2 (Continued)
| South Africa |  |  |
| --- | --- | --- |
| (Lin et al., 2022)<sup>37</sup> |  |  |
|  | N | Value (%) |
| Total Number of Patients | 300 | 100.00% |
| COVID-19 Strain(s) |  |  |
| Omicron | 300 | 100.00% |
| Delta | * | * |
| Alpha | * | * |
| Sex |  |  |
| Female | 132 | 44.00% |
| *Omicron* | 132 | 44.00% |
| *Delta* | * | * |
| *Alpha* | * | * |
| Male | 168 | 56.00% |
| *Omicron* | 168 | 56.00% |
| *Delta* | * | * |
| *Alpha* | * | * |

Note. \*=Data not reported

Table 2 (Continued)
|  | **South Africa** |  | **China** |  | **Japan** |  |
| --- | --- | --- | --- | --- | --- | --- |
|  | **(Abdullah et al., 2022)<sup>28</sup>** |  | **(Bi et al., 2022)<sup>29</sup>** |  | **(Ito et al., 2022)<sup>30</sup>** |  |
|  | N | Value (%) | N | Value (%) | N | Value (%) |
| **Age** |  |  |  |  |  |  |
| 60+ | 1,521 | 34.3 | * | * | * | * |
| *Omicron* | 1,521 | 34.3 | * | * | * | * |
| *Delta* | * | * | * | * | * | * |
| *Alpha* | * | * | * | * | * | * |
| <60 | 2,877 | 65.0 | * | * | * | * |
| *Omicron* | 2,877 | 65.0 | * | * | * | * |
| *Delta* | * | * | * | * | * | * |
| *Alpha* | * | * | * | * | * | * |
| **Vaccination** |  |  |  |  |  |  |
| Zero Doses | * | * | * | * | 80 | 34.6 |
| *Omicron* | * | * | * | * | 80 | 34.6 |
| *Delta* | * | * | * | * | * | * |
| *Alpha* | * | * | * | * | * | * |
| 1+ Doses | * | * | * | * | 151 | 65.4 |
| *Omicron* | * | * | * | * | 151 | 65.4 |
| *Delta* | * | * | * | * | * | * |
| *Alpha* | * | * | * | * | * | * |

Note. \*=Data not reported

Table 2 (Continued)
|  | **Canada** |  | **Bahrain** |  | **Japan** |  |
| --- | --- | --- | --- | --- | --- | --- |
|  | **(Ulloa et al., 2022)<sup>31</sup>** |  | **(Kumar et al., 2022)<sup>32</sup>** |  | **(Miyashita et al., 2022)<sup>33</sup>** |  |
|  | N | Value (%) | N | Value (%) | N | Value (%) |
| **Age** |  |  |  |  |  |  |
| 60+ | * | * | * | * | 198 | 59.1 |
| *Omicron* | * | * | * | * | * | * |
| *Delta* | * | * | * | * | * | * |
| *Alpha* | * | * | * | * | 198 | 59.1 |
| <60 | * | * | * | * | 137 | 40.9 |
| *Omicron* | * | * | * | * | * | * |
| *Delta* | * | * | * | * | * | * |
| *Alpha* | * | * | * | * | 137 | 40.9 |
| **Vaccination** |  |  |  |  |  |  |
| Zero Doses | 23,028 | 29.5 | 512 | 35.9 | * | * |
| *Omicron* | 7,634 | 9.8 | 293 | 20.5 | * | * |
| *Delta* | 15,394 | 19.7 | 219 | 15.3 | * | * |
| *Alpha* | * | * | * | * | * | * |
| 1+ Doses | 55,138 | 70.5 | 916 | 64.1 | * | * |
| *Omicron* | 33,582 | 43.0 | * | * | * | * |
| *Delta* | 21,556 | 27.6 | 343 | 24.0 | * | * |
| *Alpha* | * | * | 573 | 40.1 | * | * |

Note. \*=Data not reported

Table 2 (Continued)
|  | Spain (Martínez-García et al., 2021)<sup>34</sup> |  | England (Twohig et al., 2022)<sup>35</sup> |  | Singapore (Ong et al., 2022)<sup>36</sup> |  |
| --- | --- | --- | --- | --- | --- | --- |
|  | N | Value (%) | N | Value (%) | N | Value (%) |
| Age |  |  |  |  |  |  |
| 60+ | * | * | 2,718 | 6.3 | * | * |
| *Omicron* | * | * | * | * | * | * |
| *Delta* | * | * | 437 | 1.0 | * | * |
| *Alpha* | * | * | 2,281 | 5.3 | * | * |
| <60 | * | * | 22,707 | 52.4 | * | * |
| *Omicron* | * | * | * | * | * | * |
| *Delta* | * | * | 8,245 | 19.0 | * | * |
| *Alpha* | * | * | 14,462 | 33.4 | * | * |
| Vaccination |  |  |  |  |  |  |
| Zero Doses | * | * | 32,078 | 74.0 | 106 | 85.5 |
| *Omicron* | * | * | * | * | * | * |
| *Delta* | * | * | 6,255 | 14.4 | 49 | 39.5 |
| *Alpha* | * | * | 25,823 | 59.6 | 57 | 46.0 |
| 1+ Doses | * | * | 11,260 | 26.0 | 18 | 14.5 |
| *Omicron* | * | * | * | * | * | * |
| *Delta* | * | * | 2,427 | 5.6 | 18 | 14.5 |
| *Alpha* | * | * | 8,833 | 20.4 | 0 | 0.0 |

Note. \*=Data not reported

Table 2 (Continued)
| South Africa |  |  |
| --- | --- | --- |
| (Lin et al., 2022)<sup>37</sup> |  |  |
|  | N | Value (%) |
| Age |  |  |
| 60+ | * | * |
| *Omicron* | * | * |
| *Delta* | * | * |
| *Alpha* | * | * |
| <60 | * | * |
| *Omicron* | * | * |
| *Delta* | * | * |
| *Alpha* | * | * |
| Vaccination |  |  |
| Zero Doses | 62 | 20.7 |
| *Omicron* | 62 | 20.7 |
| *Delta* | * | * |
| *Alpha* | * | * |
| 1+ Doses | 238 | 79.3 |
| *Omicron* | 238 | 79.3 |
| *Delta* | * | * |
| *Alpha* | * | * |

Note. \*=Data not reported

Table 2 (Continued)
|  | **South Africa** |  | **China** |  | **Japan** |  |
| --- | --- | --- | --- | --- | --- | --- |
|  | **(Abdullah et al., 2022)<sup>28</sup>** |  | **(Bi et al., 2022)<sup>29</sup>** |  | **(Ito et al., 2022)<sup>30</sup>** |  |
|  | N | Value (%) | N | Value (%) | N | Value (%) |
| Signs |  |  |  |  |  |  |
| Fever | * | * | 61 | 93.8 | 79 | 34.2 |
| *Omicron* | * | * | 61 | 93.8 | 79 | 34.2 |
| *Delta* | * | * | * | * | * | * |
| *Alpha* | * | * | * | * | * | * |
| Pneumonia | * | * | * | * | * | * |
| *Omicron* | * | * | * | * | * | * |
| *Delta* | * | * | * | * | * | * |
| *Alpha* | * | * | * | * | * | * |

Note. \*=Data not reported

Table 2 (Continued)
|  | **Canada** |  | **Bahrain** |  | **Japan** |  |
| --- | --- | --- | --- | --- | --- | --- |
|  | **(Ulloa et al., 2022)<sup>31</sup>** |  | **(Kumar et al., 2022)<sup>32</sup>** |  | **(Miyashita et al., 2022)<sup>33</sup>** |  |
|  | N | Value (%) | N | Value (%) | N | Value (%) |
| **Signs** |  |  |  |  |  |  |
| Fever | * | * | * | * | 295 | 88.1 |
| *Omicron* | * | * | * | * | * | * |
| *Delta* | * | * | * | * | * | * |
| *Alpha* | * | * | * | * | * | * |
| Pneumonia | * | * | * | * | * | * |
| *Omicron* | * | * | * | * | * | * |
| *Delta* | * | * | * | * | * | * |
| *Alpha* | * | * | * | * | * | * |

*Note.* \*=Data not reported

Table 2 (Continued)
|  | Spain (Martínez-García et al., 2021)<sup>34</sup> |  | England (Twohig et al., 2022)<sup>35</sup> |  | Singapore (Ong et al., 2022)<sup>36</sup> |  |
| --- | --- | --- | --- | --- | --- | --- |
|  | N | Value (%) | N | Value (%) | N | Value (%) |
| Signs |  |  |  |  |  |  |
| Fever | * | * | * | * | 81 | 65.3 |
| *Omicron* | * | * | * | * | * | * |
| *Delta* | * | * | * | * | 48 | 38.7 |
| *Alpha* | * | * | * | * | 33 | 26.6 |
| Pneumonia | * | * | * | * | 42 | 33.9 |
| *Omicron* | * | * | * | * | * | * |
| *Delta* | * | * | * | * | 33 | 26.6 |
| *Alpha* | * | * | * | * | 9 | 7.3 |

Note. \*=Data not reported

Table 2 (Continued)
| South Africa |  |  |
| --- | --- | --- |
| (Lin et al., 2022)<sup>37</sup> |  |  |
|  | N | Value (%) |
| Signs |  |  |
| Fever | * | * |
| *Omicron* | * | * |
| *Delta* | * | * |
| *Alpha* | * | * |
| Pneumonia | * | * |
| *Omicron* | * | * |
| *Delta* | * | * |
| *Alpha* | * | * |

Note. \*=Data not reported

The results of the *X*^2^ tests suggest that most bivariate associations between demographic factors and COVID-19 strains are significant (Table 3). Vaccination is associated with testing positive for each of the COVID-19 strains of interest, irrespective of geographic setting. n most geographical regions, a significant statistical association has been observed between age, sex, vaccination status, and testing positive for various strains of COVID-19. However, it is important to note that this association does not hold true universally across all locations. The sampled studies from Canada and Singapore show no significant relationship between sex and testing positive for Delta and Omicron or Delta and Alpha when pooled in the multi-strain category. The COVID-19 strains, Delta and Alpha (when pooled in the multi-strain category), didn’t show a significant association with sex in Singapore (*X*^2^ = 1.42, p-value = 0.5). The COVID-19 strains, Delta and Omicron (when pooled in the multi-strain category), did not show a significant association with sex in Canada (*X*^2^ = 1.41, p-value = 0.495).

**Table 3.** Relative odds ratio and chi-squares association of contracting a certain COVID-19 strain based on potential risk factor (age, sex, and vaccination)

|  | **South Africa** |  | **China** |  | **Japan** |  |
| --- | --- | --- | --- | --- | --- | --- |
|  | **(Abdullah et al., 2022)<sup>28</sup>** |  | **(Bi et al., 2022)<sup>29</sup>** |  | **(Ito et al., 2022)<sup>30</sup>** |  |
| | $\chi^2$ | OR | $\chi^2$ | OR | $\chi^2$ | OR |
|  | (P-Value) | (95% CI) | (P-Value) | (95% CI) | (P-Value) | (95% CI) |
| Sex | * | * | 16.5<br>(0.00026) | * | 58.3<br>(2.24E-13) | * |
| Age | 1440<br>(<2.2E-16) | * | * | * | * | * |
| Vaccination | * | * | * | * | 74.1<br>(<2.2E-16) | * |

*Note.* <sup>1</sup> = Result is not significant (P-value > 0.05)

Table 3 (Continued)
|  | **Canada** |  | **Bahrain** |  | **Japan** |  |
| --- | --- | --- | --- | --- | --- | --- |
|  | **(Ulloa et al., 2022)<sup>31</sup>** |  | **(Kumar et al., 2022)<sup>32</sup>** |  | **(Miyashita et al., 2022)<sup>33</sup>** |  |
| | $\chi^2$ (P-Value) | OR (95% CI) | $\chi^2$ (P-Value) | OR (95% CI) | $\chi^2$ (P-Value) | OR (95% CI) |
| Sex | 1.41<br>(0.495)<sup>1</sup> | 0.98<br>(0.96–1.01)<sup>1</sup> | 33.7<br>(4.70E-08) | 0.49<br>(0.39–0.63) | 115<br>(<2.2E-16) | * |
| Age | * | * | * | * | 92.1<br>(<2.2E-16) | * |
| Vaccination | 5020<br>(<2.2E-16) | 3.14<br>(3.04–3.24) | 52.0<br>(5.05E-12) | 0.45<br>(0.36–0.56) | * | * |

*Note.* <sup>1</sup> = Result is not significant (P-value > 0.05)

Table 3 (Continued)
|  | **Spain** |  | **England** |  | **Singapore** |  |
| --- | --- | --- | --- | --- | --- | --- |
|  | **(Martínez-García et al., 2021)<sup>34</sup>** |  | **(Twohig et al., 2022)<sup>35</sup>** |  | **(Ong et al., 2022)<sup>36</sup>** |  |
| | $\chi^2$ | OR | $\chi^2$ | OR | $\chi^2$ | OR |
|  | (P-Value) | (95% CI) | (P-Value) | (95% CI) | (P-Value) | (95% CI) |
| Sex | 117<br>($<2.2\text{E-}16$) | * | 21.0<br>($2.79\text{E-}05$) | 0.90<br>(0.85–0.94) | 1.42<br>(0.492)<sup>1</sup> | 1.54<br>(0.75–1.65)<sup>1</sup> |
| Age | * | * | 442<br>($<2.2\text{E-}16$) | 2.97<br>(2.68–3.10) | 17.9<br>($1.29\text{E-}04$) | * |
| Vaccination | * | * | 22.0<br>($1.70\text{E-}05$) | 1.13<br>(1.08–1.20) | * | * |

Note. <sup>1</sup> = Result is not significant (P-value > 0.05)

Table 3 (Continued)
| **South Africa** |  |  |
| --- | --- | --- |
| **(Lin et al., 2022)<sup>37</sup>** |  |  |
| | $\chi^2$ | OR |
|  | (P-Value) | (95% CI) |
| Sex | 78.2<br>($<2.2\text{E-}16$) | * |
| Age | * | * |
| Vaccination | 152<br>($<2.2\text{E-}16$) | * |

*Note.* <sup>1</sup> = Result is not significant (P-value > 0.05)

To understand the strength and direction of the bivariate associations between demographic factors and COVID-19 infection, we also calculated odds ratios with 95% confidence intervals. Results of the odds ratio test with the potential risk factors of COVID-19 infection as the dependent variable are shown in Figure 2. Though not statistically significant, we found males had higher odds than females of testing positive for Alpha during the Alpha-Delta wave in Singapore (OR = 1.54 (0.750 - 3.19), p-value = 0.241); the odds of a female having Delta are similar to the odds of a male having Delta in a Delta-Omicron wave in Canada, even though it is not statistically significant. (OR = 0.98 (0.96–1.01), p-value = 0.236). For sex, males have a higher odds than females of testing positive for Alpha during an Alpha-Delta wave in Bahrain and England. Additionally, the odds of an individual who is 60 years or older having Alpha are 2.97 times the odds of an individual who is less than 60 years in an Alpha-Delta wave (OR = 2.97 (2.68–3.10), p-value = 0). For vaccination, there are contradicting results about if unvaccinated or vaccinated individuals have a higher odds of having Alpha in an Alpha-Delta wave. Furthermore, the largest odds ratio was observed for vaccination and having Alpha in a Delta-Omicron wave.

**Figure 2.**
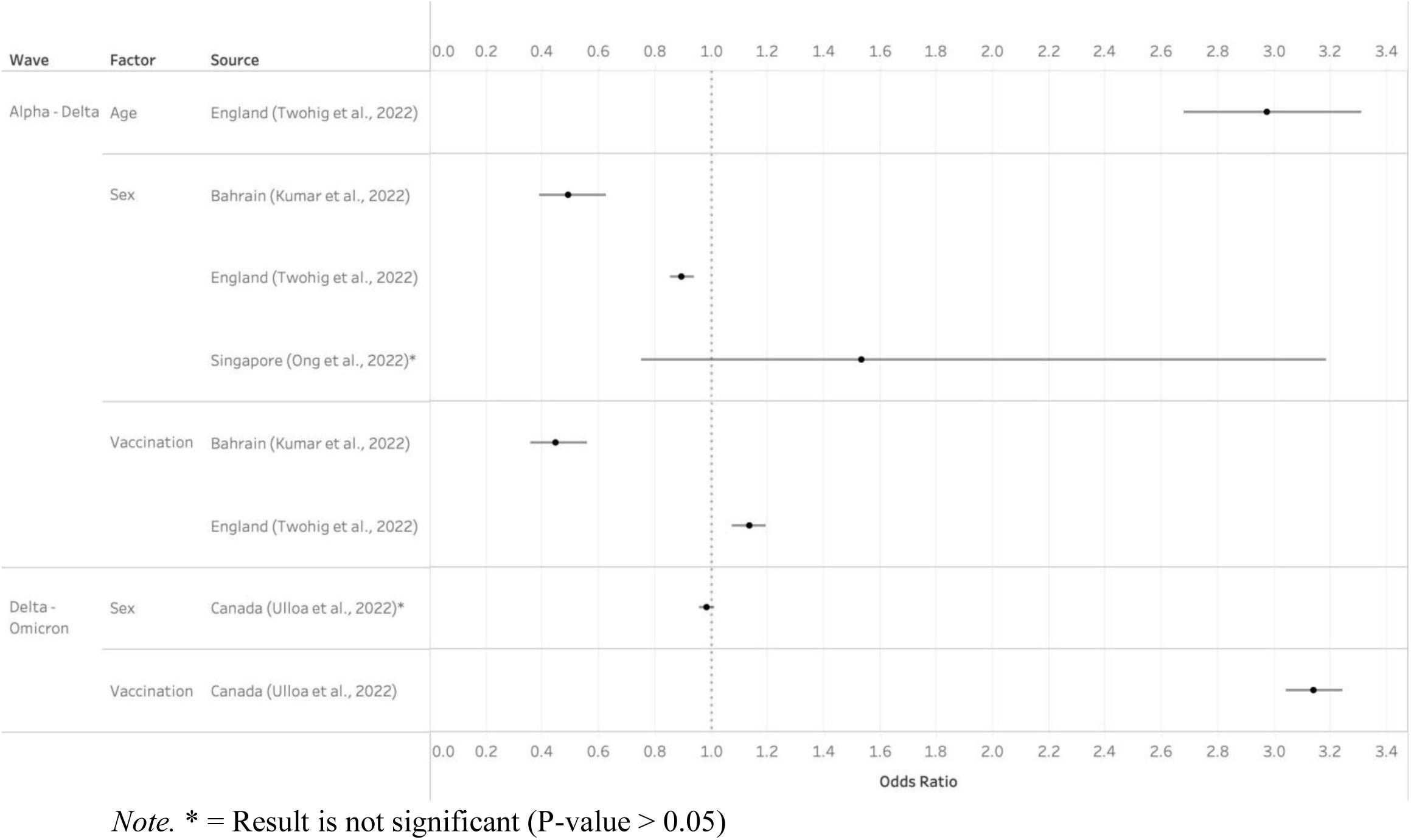
Forest plot of odds ratios and 95% confidence interval based on the wave, factor, and source.

## Discussion

COVID-19 strains have impacted regions across the world in various ways. It is necessary to understand the odds of contracting past COVID-19 strains to understand new COVID-19 strains better. This scenario facilitates the improved diagnosis of COVID-19 patients by identifying the specific strain they are infected with. Sex, age, and vaccination are the three characteristics of a COVID-19 patient that are well recorded, making it critical and reliable to understand the odds of contracting of COVID-19 strains based on these factors.

The latest studies on the association between getting infected by COVID-19 and certain demographics tend to focus on age, sex, and vaccination.^9–12^ We observed a statistically significant association between age and contracting COVID-19 regardless of the strain in all the regions observed in the present study. The current study’s findings align with previous research conducted in Europe, which has identified an association between age and contracting COVID-19.^4,13^ The association between age and contracting COVID-19 (for any of the three strains in the present study) could depend on physiologic changes in the immune system such as immunosenescence due to aging.^14,15^ In the present study, a statistically significant association was identified between being vaccinated and getting infected with COVID-19, irrespective of the viral strain, in the regions studied. The findings of the present study are consistent with prior research, which has established a relationship between vaccine coverage and transmission of infection of COVID-19 variants across different regions globally.^8,16^ The observed association between vaccination and COVID-19 infection is likely explained by the vaccine’s novel mechanism of action and overall immunology response, which is independent of geography.^17–19^ Males tend to have a greater risk of COVID-19 infection than females.^13,20^ Although in this study, sex didn’t always have a statistically significant association with the contracted COVID- 19 strain, which aligns with the idea that males and females are at an equal risk of COVID-19 infection.^21^ The present study’s findings wasn’t a significant association of sex and testing positive for specific COVID-19 strains in Singapore and Canada, but the rest of the places included in the study had a significant association with sex and contracted COVID-19 strain. The present study’s result can be interpreted as the nations that have sex equality in health don’t have a significant association between sex and COVID-19 strains.

Prior research has established that males tend to have be at higher risk of COVID-19 infection than females.^13,20^ However, in the present study, the odds of contracting Delta among females are similar to the odds of males contracting Delta in a Delta-Omicron wave; nevertheless, males have lower odds of contracting Alpha than females in the Alpha-Delta wave. This is contradicting with the theory that female COVID-19 patients have higher levels of estrogen than male COVID-19 patients which may decrease the odds of severe infection of COVID-19 since estrogen has a role in the regulation of the immune response by interfering with B cell function, resulting in a Th2 response, while testosterone suppresses the natural killer cells’ response.^22,23^

The administration of COVID-19 vaccinations serves as a crucial mechanism in minimizing the transmission and prevalence of the virus.^8,16,24^ In the Alpha-Delta wave, vaccinated people have greater odds of getting Alpha in Bahrain, but unvaccinated people have slightly greater odds of contracting Alpha in England. In the Delta-Omicron wave, unvaccinated individuals have a much higher odds of contracting Delta than the vaccinated individuals in Canada. These contradicting results can be best explained by countries like England and Canada having a plan to increase vaccination administration or knowledge of vaccine administration unlike countries like Bahrain in early 2021.^25–27^ Also, the vaccines used in Bahrain may not be approved - unlike the vaccines commonly used in England or Canada - so the vaccination in Bahrain may have not been as effective as those in Canada or England.

Aging has been known to decrease immunity since there are fewer immune cells in the body starting from around age 60.^4,14^ In the present study, there is a higher odds of individuals 60 years and older contracting Alpha than individuals who are less than 60 years old in an Alpha-Delta wave. This can be attributed to the effect of fewer B and T cells that move from primary to secondary lymphoid organs in older individuals than in younger individuals.^14,23^

This study affirms that demographic factors such as sex and age and vaccination status have a significant association with COVID-19 strains depending on the location of the individual.

There are important limitations of the present meta-analysis. First, this study isn’t a true meta-analysis; this study’s purpose is to understand how COVID-19 is associated with factors such as sex, age, and vaccination. The next step would be to conduct a true meta-analysis once more studies are published on all COVID-19 strains. Second, bivariate associations don’t account for confounding and effect modification when making public health inferences. For example, women may be more vulnerable to risk of COVID-19 infection than male in Canada because of factors such as pregnancy, or underlying diseases which weren’t accounted for in this study. Future studies should model the associations in a multivariate context to account for this. Finally, some sources have small sample sizes, preventing the data from being properly generalized to a larger population.

Despite studies being published with data on the demographics and vaccination of COVID-19 patients, there aren’t many studies analyzing the data from sources based on different regions throughout the world. The findings from the present study provide meta-analytic estimates on the odds of getting infection by COVID-19 strain of interest based on demographic subgroups and vaccination. These results can inform the healthcare industry by identifying groups that are at risk of getting infected by certain COVID-19 strains and support the idea that the risk of infection by certain COVID-19 strains varies based on the location of the individual.

## Data Availability

All data produced are available online at PubMed and the Incidence and Prevalence Database (IPD).

## Acknowledgements

The authors thank Dr. Hamsa Subramaniam for her guidance and support as an advisor for this study.

## Contributors

Concept and design: DK, MR

Acquisition, analysis, or interpretation of data: DK, SM

Drafting of the manuscript: DK

Critical revision of the manuscript for important intellectual content: All authors

Statistical analysis: DK

All the authors contributed to the writing of the manuscript. All the authors agreed with the results and conclusions of the manuscript. All authors have read, and confirm that they meet, ICMJE criteria for authorship.

